# Genomic and clinical epidemiology of SARS-CoV-2 in coastal Kenya: Insights into variant circulation, reinfection, and multiple lineage importations during a post-pandemic wave

**DOI:** 10.1101/2025.03.26.25324476

**Authors:** Arnold W. Lambisia, Esther N. Katama, Edidah Moraa, John M. Mwita, Katherine Gallagher, Martin Mutunga, Emily Nyale, Joan Omungala, Mike Mwanga, Nickson Murunga, Joyce Nyiro, James Nyagwange, Charles Sande, Philip Bejon, George Githinji, Simon Dellicour, My V.T. Phan, Matthew Cotten, L. Isabella Ochola-Oyier, Edward C. Holmes, Charles N. Agoti

**Affiliations:** Kenya Medical Research Institute (KEMRI) - Wellcome Trust Research Programme (KWTRP), Kilifi, Kenya; Department of Infectious Diseases Epidemiology, London School of Hygiene and Tropical Medicine, Keppel Street, London, United Kingdom; University of Oxford, Oxford, United Kingdom; Pwani University, Kilifi, Kenya; Spatial Epidemiology Lab (SpELL), Université Libre de Bruxelles, Bruxelles, Belgium; Department of Microbiology, Immunology and Transplantation, Rega Institute & Laboratory for Clinical and Epidemiological Virology, KU Leuven, University of Leuven, Leuven, Belgium; College of Health Solutions, Arizona State University, Phoenix, Arizona, USA; Complex Adaptive System Initiative, Arizona State University, Scottsdale, Arizona, USA; School of Medical Sciences, University of Sydney, Australia

**Keywords:** COVID-19, SARS-CoV-2, Omicron, XBB.2.3, JN.1, Reinfections, Coastal Kenya

## Abstract

Between November 2023 and March 2024, coastal Kenya experienced a new wave of severe acute respiratory syndrome coronavirus 2 (SARS-CoV-2) infections detected through our continued genomic surveillance. Herein, we report the clinical and genomic epidemiology of SARS-CoV-2 infections from 179 individuals (total 185 positive samples) residing in the Kilifi Health and Demographic Surveillance (KHDSS) area (∼900 km^2^). Sixteen SARS-CoV-2 lineages within three sub-variants (XBB.2.3-like (58.4%), JN.1-like (40.5%) and XBB.1-like (1.1%)) were identified. Symptomatic infection rate was estimated at 16.0% (95% CI 11.1%-23.9%) based on community testing regardless of symptom status, and did not differ across the sub-variants (*p =* 0.13). The most common infection symptoms in community cases were cough (49.2%), fever (27.0%), sore throat (7.3%), headache (6.9%), and difficulty in breathing (5.5%) and one case succumbed to the infection. Genomic analysis of the virus from serial positives samples confirmed repeat infections among five participants under follow-up (median interval 21 days, range 16-95 days); in four participants, the same virus lineage was responsible in both the first and second infections, while one participant had a different lineage in the second infection compared to the first. Phylogenetic analysis including >18,000 contemporaneous global sequences estimated that at least 38 independent virus introduction events occurred into the KHDSS area during the wave, the majority likely originating in North America and Europe. Our study highlights coastal Kenya, like most other localities, continues to face new SARS-CoV-2 infection waves characterized by the circulation of new variants, multiple lineage importations and reinfections. Locally the virus may circulate unrecognized as most infections are asymptomatic in part due to high population immunity after several waves of infection. Our findings highlight the need for sustained SARS-CoV-2 surveillance to inform appropriate public health responses such as scheduled vaccination for risk populations.

**Author summary:** Severe acute respiratory syndrome coronavirus 2 (SARS-CoV-2) has transitioned to an endemic respiratory pathogen causing seasonal outbreaks. We examined the epidemiological and genomic patterns of a wave of SARS-CoV-2 infections in coastal Kenya that occurred between November 2023 and March 2024. By analyzing genetic and epidemiological data from positive cases in Kilifi, we inferred the origins of the new strains, documented repeat infections and the virus’ ongoing evolution. Our data revealed several variants circulating in the community, indicating multiple new virus introductions probably before and during local outbreaks. Many infected individuals were asymptomatic, highlighting unnoticed transmission within the population. Despite low vaccination rates among the cases (∼7.0%), high population immunity has previously been reported locally. The common symptoms among those who were symptomatic included cough, fever, and sore throat. A few participants experienced repeat infections during the wave, often involving closely related strains. The virus lineages detected were most closely related to those sampled in Europe. Our findings emphasize that, despite the end of the emergency phase, SARS-CoV-2 remains a significant public health issue, necessitating ongoing monitoring and responsive measures e.g. target vaccinations, masking and good hygiene practices to protect those at risk of severe infection.

## Introduction

During the coronavirus disease 2019 (COVID-19) pandemic (2020-2023), there was a globally unprecedented levels of virus genomic surveillance with >14 million partial or complete genomes of SARS-CoV-2 (the causative agent of COVID-19) ^1^. This enabled detailed investigation into the molecular epidemiology of SARS-CoV-2, in turn revealing the emergence of variants of concern (VOC), modes of evolution (including single nucleotide polymorphisms (SNPs), recombination, sequence deletions, etc.) and the pathways of virus spread across different scales of observation, information which together contributed to the rational design and key updates of control tools including vaccine strain composition for booster doses^2^.

When the World Health Organisation (WHO) declared an end to the public health emergency of international concern status (PHEIC) of COVID-19 in May 2023 ^3^, SARS-CoV-2 genomic surveillance substantially decreased, albeit its clearly demonstrated value ^4,5^. This has led to a paucity of systematically collected genomic and epidemiological data for recently emerged SARS-CoV-2 variants/lineages e.g., JN.1, XBB.2.3, LF.8, XEC, especially in low- and middle-income country settings (LMICs). In the meantime, waves of SARS-CoV-2 infections continue to appear, especially with the emergence of novel variants associated with increased transmissibility, virulence, and/or immune escape ^67^.

Kenya, a lower-middle income country in East Africa with a population of ∼50 million people, recorded its first COVID-19 case on March 13^th^, 2020, ^8^. COVID-19 vaccination started in the country in March 2021 and as of November 2022, approximately 11.1% of the Kenyan population had received ≥ 1 dose of COVID-19 vaccine ^9^. Since March 2020, the country has consistently recorded 1-2 waves of SARS-CoV-2 infections annually, with the coastal region of Kenya reporting seven waves of SARS-CoV-2 infection by the end of May 2023 ^10–13^. The waves were dominated by: B.1* (wave peak in July 2020), B.1* (wave peak in November 2020) ^13^, B.1.1.7 (Alpha VOC) and B.1.351 (Beta VOC; March-April 2021), B.1.617.2* (Delta VOC; July 2021) ^14^, BA.1* (Omicron VOC; January 2022), BA.4/5* (Omicron; June 2022), BQ.1* (Omicron; November 2022), and FY.4* (Omicron, alias XBB.1.22.1.4; April 2023) ^15^.

While there is substantially reduced SARS-CoV-2 genomic surveillance, in Kenya, continuous genomic surveillance had generated at least 13,816 SARS-CoV-2 partial or complete genome sequences available in the Global Initiative on Sharing All Influenza Data (GISAID) database as of October 6, 2024, enabling a comprehensive tracking of the epidemic at a national scale ^11–14,16^. By the end of 2022, more than 70% of Kilifi residents (a rural population in coastal Kenya) and 90% of Nairobi residents (an urban population in Kenya’s capital city) were seropositive for SARS-CoV-2 anti-S IgG ^9^. Despite this apparent high infection rates, Kenya like most other sub-Saharan Africa countries, experienced a relatively mild CoVID-19 disease burden, with >80% of the infections asymptomatic. However, data to support this conclusion are limited to a few community studies ^17,18^. As new waves of infection appear in the region, genomic surveillance remains an important tool to identify the variants involved, their evolution, and their impact on current control measures such as vaccines.

The KEMRI-Wellcome Trust Research Programme (KWTRP) runs the Kilifi Health and Demographic Surveillance System (KHDSS; ∼300,000 residents in 900 km^2^ area) ^19^. Within the KHDSS, KWTRP currently runs three surveillance platforms for monitoring respiratory disease epidemiology: community (weekly homestead visits) surveillance, outpatient (five health facilities) surveillance, and inpatient paediatric (<5-year-olds) pneumonia surveillance (Kilifi County Hospital, **Fig 1**) ^20,21^. The patterns of repeat infections during new community waves of SARS-CoV-2 transmission have not been reported anywhere in Africa outside of South Africa ^22^. Here, we describe the epidemiological patterns from the analyses of SARS-CoV-2 genomic data collected at the KHDSS during the wave of SARS-CoV-2 infections that occurred between November 2023 and March 2024 in coastal Kenya (eighth wave).

**Fig 1:**
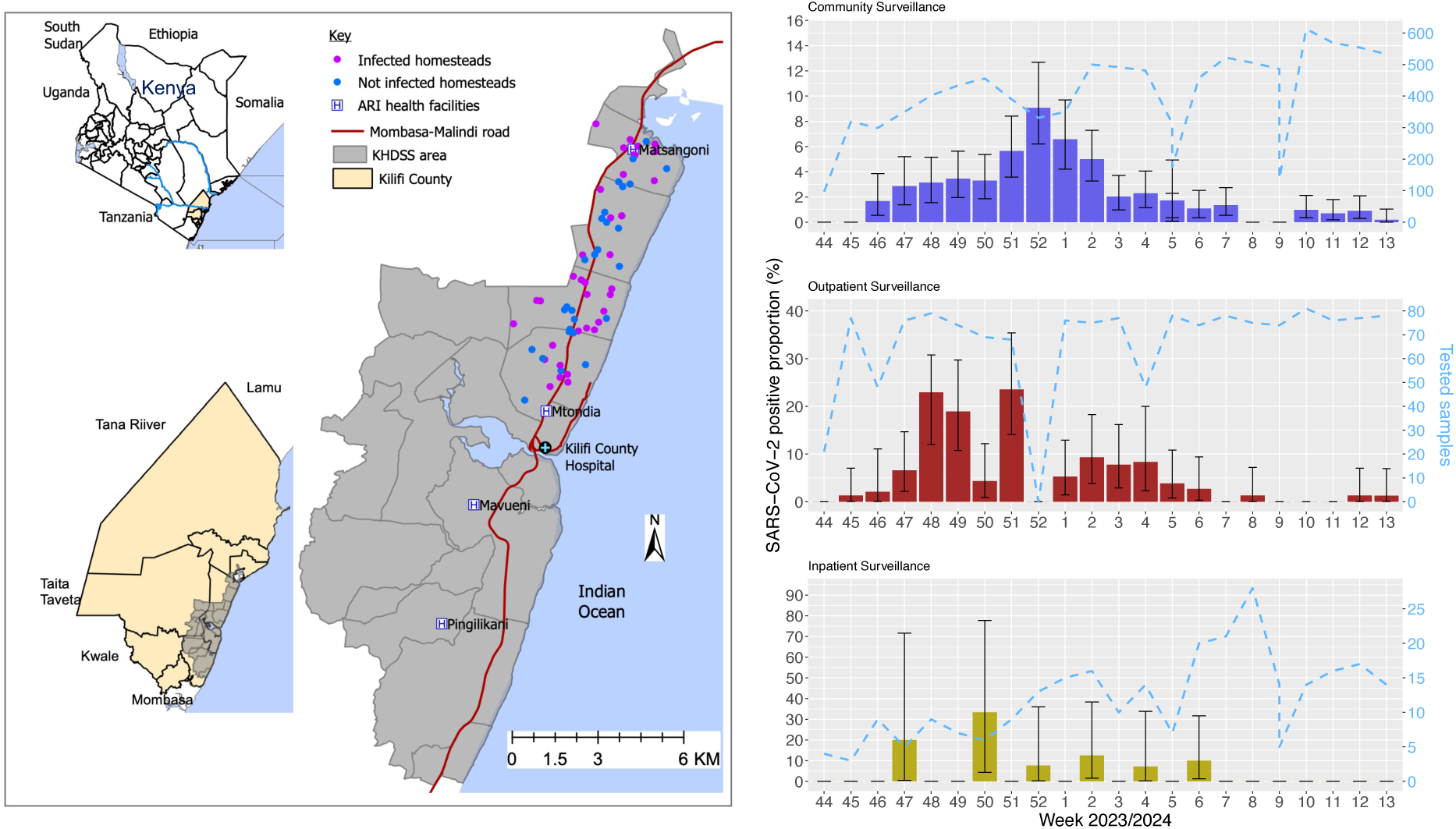
Study population and surveillance platforms. **(Left panel)** A map of the study area showing Kilifi Health and Demographic Surveillance System (KHDSS), Kilifi, Kenya (the big map) and its location within Kilifi County and Kenya. The pink and blue dots show the distribution of the recruited homesteads in the community surveillance arm. The location of the five health facilities for inpatient (KCH) and/or outpatient surveillance is indicated. **(Right panel)** Temporal trends of SARS-CoV-2 positive samples starting October 2023 to March 2024 in Kilifi, Kenya. Specifically, we report weekly SARS-CoV-2 detection in KHDSS area between October 2023 and March 2024 across the three surveillance platforms: community, outpatient, and inpatient. The dashed line shows the total number of samples tested per week (secondary y-axis) across the different platforms.

## Results

### SARS-CoV-2 infections in the KHDSS – November 2023 to March 2024

The 2023-24 SARS-CoV-2 wave of infections in the KHDSS began in the second week of November and peaked in the third week of December 2023, after which infection numbers gradually reduced toward the end of March 2024 as evident across the three surveillance platforms (**Fig 1A-C**). This represents the eighth wave of SARS-CoV-2 in the KHDSS. Between August 31, 2023, and March 31,2024, a total of 13,847 nasopharyngeal/oropharyngeal (NP/OP) swabs samples from 3,150 individuals were screened for SARS-CoV-2 by a quantitative PCR (qPCR) targeting the envelope (E) gene. Of these, 308 samples from 235 individuals were found positive (cycle threshold (Ct) <35.0). These were collected from 138 individuals enrolled in the community surveillance platform, 88 in the outpatient platform, and 9 in the inpatient platform.

During the peak month of the wave (December 2023), the sample positivity rate was 5.0% (95% CI: 4.0-6.2%) in the community, 15.2% (95% CI: 11.0-20.7%) in outpatients and 7.6% (95% CI: 2.0-21.9%) in the inpatients (**Fig 1A-C**). There were slightly more female positive cases among the identified infections than males (*n* = 136, 56.4%) and the median age among positive cases was 14.0 years (interquartile range 3.0 – 30.0 years; **Table 1**). Seven participants enrolled in the community study were suspected of experiencing a repeat infection episode during the outbreak (defined as positive NP/OP samples separated by 14 or more days, with intervening negative tests; **S1 Fig**). Among these seven suspected reinfection cases, six were females. Four of the cases involved individuals aged 0 to 1 year, one was in the 2 to 4-year age group, one was in the 5 to 9-year age group, and one was in the 20 to 39-year age group. One case reported having asthma as a comorbidity at the time of recruitment(**Table 2**).

**Table 1.** Baseline characteristics of SARS-CoV-2 cases identified in three KHDSS surveillance platforms between November 2023 and March 2024.

|  | Community | Outpatient | Inpatient | Total |
| --- | --- | --- | --- | --- |
| Total samples analyzed | 11,355 | 2,151 | 341 | 13,847 |
| Total participants | 658 | 2,151 | 341 | 3,150 |
| Positive samples | 211 | 88 | 9 | 308 |
| Total cases positive | 144* | 88 | 9 | 241 |
| Sequencing status |  |  |  |  |
| Sequenced | 115 (79.9) | 65 (73.9) | 5 (55.6) | 185 (76.8) |
| Sex |  |  |  |  |
| Female | 52 (59.1) | 2 (22.2) | 82 (56.9) | 136 (56.4) |
| Age (Years) |  |  |  |  |
| Median (IQR) | 12 (3-30.5) | 21 (4.7-31.7) | 0 (0-0) | 14.0 (3.0-30.0) |
| Age group |  |  |  |  |
| 0-4 | 44 (30.6) | 22 (25.0) | 9 (100.0) | 75 (31.1) |
| 5-9 | 19 (13.2) | 5 (5.7) | - | 24 (10.0) |
| 10-19 | 30 (20.8) | 14 (15.9) | - | 44 (18.3) |
| 20-39 | 27 (18.8) | 30 (34.1) | - | 57 (23.7) |
| 40-64 | 17 (11.8) | 9 (10.2) | - | 26 (10.8) |
| 65+ | 7 (4.9) | 8 (9.1) | - | 15 (6.2) |
| Symptom status |  |  |  |  |
| Asymptomatic | 120 (83.3%) | 0 (0.0%) | 0 (0.0%) | 120 (49.8%) |
| Symptomatic | 24 (16.7%) | 88 (100.0%) | 9 (100.0%) | 121 (50.2%) |
| Vaccination Status |  |  |  |  |
| Yes | 10 (6.9) | 12 (13.6) | - | 22 (9.1) |
| No | 134 (93.1) | 76 (86.4) | 9 (100.0) | 219 (90.9) |

\*Note that this includes seven participants in the community study named respiratory virus reinfections (ResViRe) who had suspected repeat infection episodes.

**Table 2.**
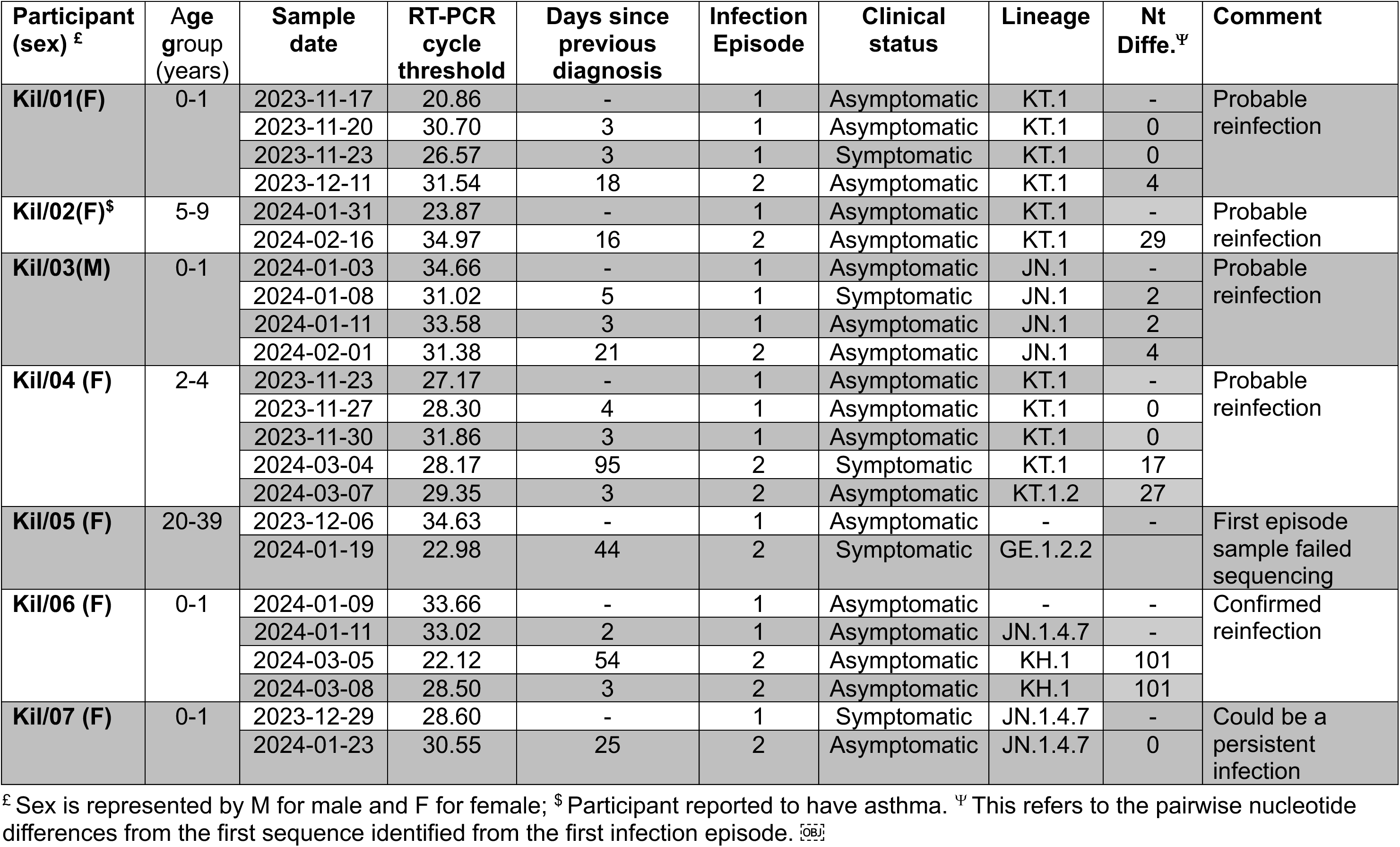
Demographic, Clinical, and Laboratory Findings for Seven Suspected Reinfection Cases from Community Surveillance Study (ResViRe)

| Participant (sex) <sup>£</sup> | Age group (years) | Sample date | RT-PCR cycle threshold | Days since previous diagnosis | Infection Episode | Clinical status | Lineage | Nt Diffe. <sup>¥</sup> | Comment |
| --- | --- | --- | --- | --- | --- | --- | --- | --- | --- |
| Kil/01(F) | 0-1 | 2023-11-17 | 20.86 | - | 1 | Asymptomatic | KT.1 | - | Probable reinfection |
|  |  | 2023-11-20 | 30.70 | 3 | 1 | Asymptomatic | KT.1 | 0 |  |
|  |  | 2023-11-23 | 26.57 | 3 | 1 | Symptomatic | KT.1 | 0 |  |
|  |  | 2023-12-11 | 31.54 | 18 | 2 | Asymptomatic | KT.1 | 4 |  |
| Kil/02(F)<sup>\$</sup> | 5-9 | 2024-01-31 | 23.87 | - | 1 | Asymptomatic | KT.1 | - | Probable reinfection |
|  |  | 2024-02-16 | 34.97 | 16 | 2 | Asymptomatic | KT.1 | 29 |  |
| Kil/03(M) | 0-1 | 2024-01-03 | 34.66 | - | 1 | Asymptomatic | JN.1 | - | Probable reinfection |
|  |  | 2024-01-08 | 31.02 | 5 | 1 | Symptomatic | JN.1 | 2 |  |
|  |  | 2024-01-11 | 33.58 | 3 | 1 | Asymptomatic | JN.1 | 2 |  |
|  |  | 2024-02-01 | 31.38 | 21 | 2 | Asymptomatic | JN.1 | 4 |  |
| Kil/04 (F) | 2-4 | 2023-11-23 | 27.17 | - | 1 | Asymptomatic | KT.1 | - | Probable reinfection |
|  |  | 2023-11-27 | 28.30 | 4 | 1 | Asymptomatic | KT.1 | 0 |  |
|  |  | 2023-11-30 | 31.86 | 3 | 1 | Asymptomatic | KT.1 | 0 |  |
|  |  | 2024-03-04 | 28.17 | 95 | 2 | Symptomatic | KT.1 | 17 |  |
|  |  | 2024-03-07 | 29.35 | 3 | 2 | Asymptomatic | KT.1.2 | 27 |  |
| Kil/05 (F) | 20-39 | 2023-12-06 | 34.63 | - | 1 | Asymptomatic | - | - | First episode sample failed sequencing |
|  |  | 2024-01-19 | 22.98 | 44 | 2 | Symptomatic | GE.1.2.2 |  |  |
| Kil/06 (F) | 0-1 | 2024-01-09 | 33.66 | - | 1 | Asymptomatic | - | - | Confirmed reinfection |
|  |  | 2024-01-11 | 33.02 | 2 | 1 | Asymptomatic | JN.1.4.7 | - |  |
|  |  | 2024-03-05 | 22.12 | 54 | 2 | Asymptomatic | KH.1 | 101 |  |
|  |  | 2024-03-08 | 28.50 | 3 | 2 | Asymptomatic | KH.1 | 101 |  |
| Kil/07 (F) | 0-1 | 2023-12-29 | 28.60 | - | 1 | Symptomatic | JN.1.4.7 | - | Could be a persistent infection |
|  |  | 2024-01-23 | 30.55 | 25 | 2 | Asymptomatic | JN.1.4.7 | 0 |  |

<sup>£</sup> Sex is represented by M for male and F for female; <sup>\$</sup> Participant reported to have asthma. <sup>Ψ</sup> This refers to the pairwise nucleotide differences from the first sequence identified from the first infection episode. <sup>[OBJ]</sup>

### Clinical presentation and viral load patterns

Cases captured by outpatient (n=88) and inpatient (n=9) surveillance were all symptomatic. Overall, only 9.1% of the cases described here were previously vaccinated (**Table 1**). Note that in Kenya, COVID-19 vaccination is recommended only for individuals aged 12 years and above, who represent the minority of the infections described in this study. All inpatient cases were discharged alive. The community surveillance collected weekly NP/OP samples regardless of symptom status. For most of the community surveillance positive cases, the infection episodes remained asymptomatic (*n* = 124, 83.2%). Overall, the common symptoms among positive cases were cough (49.2%), fever (above 37.8°C; 27.0%), sore throat (7.3%), headache (6.9%), and difficulty breathing (5.5%). One symptomatic case (< 1-year-old) from community surveillance died during an ongoing episode. Among the symptoms observed in this case were coughing, a fever (39.3°C), a runny nose, and wheezing. The patient was diagnosed with pneumonia and anemia, treated and discharged. Only two out of the seven suspected reinfection cases identified in the community surveillance were symptomatic (**Table 2; S1 Fig)**. Among the community cases, sex, age, vaccination status and infecting variant were not associated with the likelihood of being symptomatic during an infection episode (**S1 Table**).

Using qPCR Ct as a proxy for viral load, there were a statistically significant difference in the approximate viral load at sampling for participants across the three sampling platforms. Lower viral loads occurred among the community surveillance samples (Kruskal-Wallis, *p* = 0.0045; **S2 Fig**). Furthermore, among cases from the community surveillance, asymptomatic individuals appeared to have a slightly lower viral load (inferred from the higher median Ct value) compared to the symptomatic individuals, although the difference did not reach statistical significance (Wilcoxon, *p* = 0.33; **S2 Fig**). Among the suspected reinfections, the median Ct values were slightly lower for the first infection (28.6; IQR 25.5-32.0) compared to the reinfection cases (30.5; IQR 25.6-31.5), with a Wilcoxon test *p-value* of 0.9015.

### SARS-CoV-2 lineage variations of the Kilifi 2023-24 wave

Sequencing was performed for at least one sample per infection episode for community surveillance and all positive samples from both outpatient and inpatient surveillance platforms. A total of 185 qPCR positive samples from 179 individuals collected between November 14, 2023, and March 27, 2024, in nine locations of the KHDSS area yielded genomic sequences covering >70% genome; all publicly shared on GISAID database). The demographic characteristics including age, sex, location or facility, and clinical presentation among sequenced and non-sequenced cases were similar (**S2 Table**). The median cycle threshold (Ct) value (inverse viral load) for sequenced cases with ≥70% of the genome recovered was lower compared to the sequenced cases with <70 of the genome recovered (median 27.8, inter quartile range (IQR) 24.7 – 30.9) vs median 32.3 (IQR 28.7 – 34.1)) with a significant difference (*p* < 0.0001; **S2 Fig**).

The recovered genomic sequences were classified into 16 Pango lineages within three Omicron sub-variants – XBB.2.3-like (108 samples), JN.1-like (75 samples), and XBB.1-like (2 samples; **S3 Table** – with XBB.2.3-like and JN.1-like jointly dominating the wave (98.9% of all cases; **Fig 2**). Similar demographic and clinical characteristics were observed in participants infected with JN.1-like, XBB.1-like and XBB.2.3-like lineages (**S4 Table**). Seven lineages were identified within XBB.2.3-like cases; KT.1 (*n* = 56), KH.1 (*n* = 25), GE.1.2.2 (*n* = 16), GE.1.2 (*n* = 7), GS.4.1 (*n* = 2), KT.1.2, (*n* = 1) and JE.1.1.1 (*n* = 1; **Fig 2**). Equally, seven lineages were identified within the JN.1-like cases; JN.1 (*n* = 33), JN.1.1 (*n* = 1), JN.1.4 (*n* = 3), JN.1.4.7 (*n* = 30), JN.1.10 (*n* = 1), JN.1.16.3 (*n* = 1) and LE.1 (*n* = 5). Two lineages were identified in XBB.1-like cases; XBB.1.5 (*n* = 1) and XBB.1.34.1 (*n* = 1; **S2 Table**). Notably, the XBB.2.3-like virus during the wave was initially high, then declined, and subsequently increased again in Kilifi towards the end of the wave (**Fig. 2**). Genomic sequencing and typing revealed that earlier the virus belonged to the GE.1.2.1 and KT.1 lineages within the sub-variant, while later it was identified as KH.1, which is genetically distinct.

**Fig 2:**
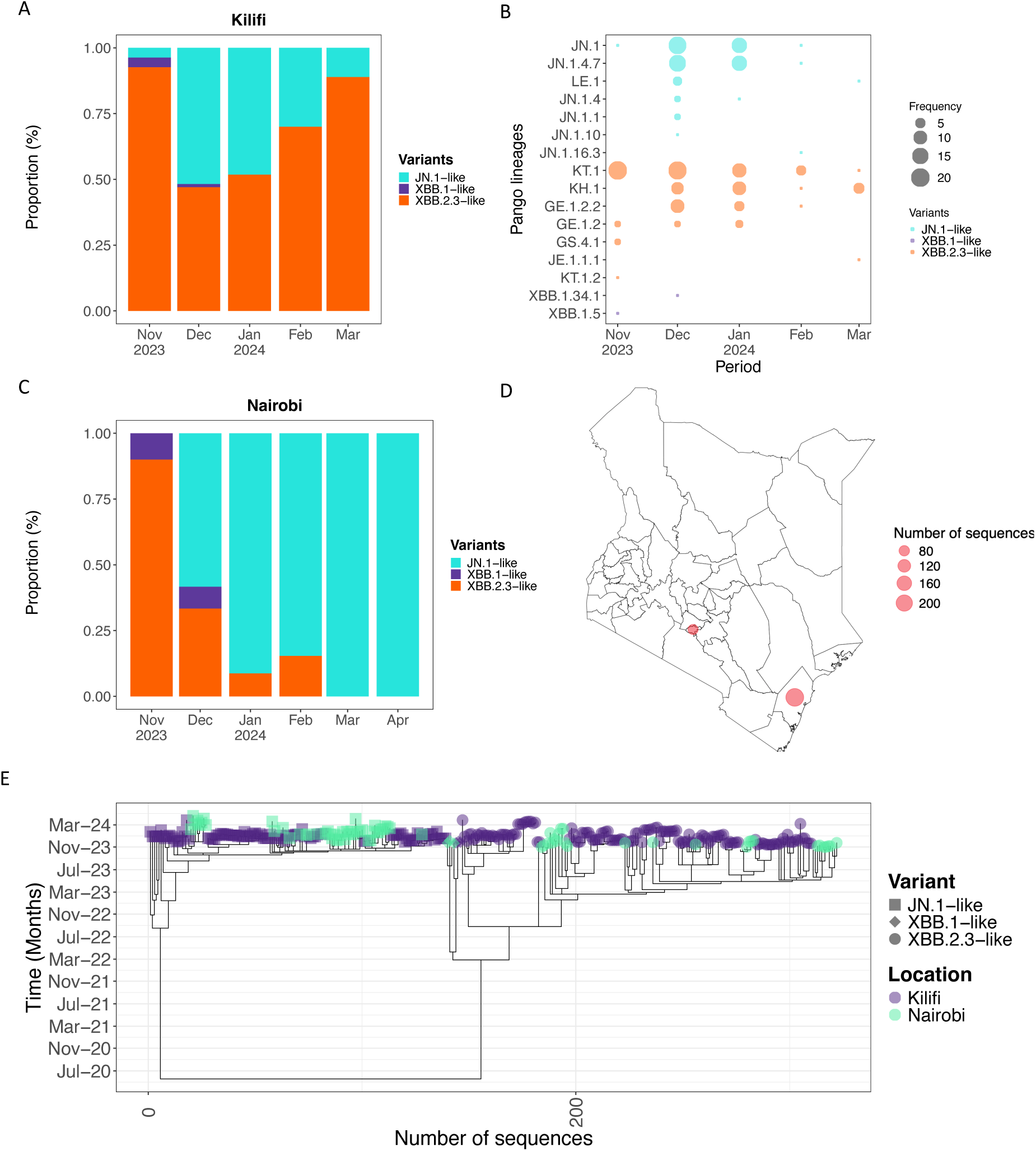
Temporal and spatial distribution of SARS-CoV-2 sub-variants/lineages in Kenya, October 2023 to March 2024. **A)** Proportionate monthly distribution of three major subvariants detected at the Kenyan Coast during the wave. **B)** Monthly numbers for the detected 16 Pango lineages during the study period **C)** Proportionate monthly distribution of three major subvariants detected in Nairobi during the wave **D)** Map of Kenya showing the two counties (Nairobi and Kilifi) that have SARS-CoV-2 whole genome data available during the study period. The size of the circles is scaled by their number of SARS-CoV-2 sequences available in GISAID. **E)** A time-resolved phylogeny showing the clustering of Kenyan sequences(n=310) collected during the study period. The different tip shapes indicate the different sub-variants detected and are colored by the county of sample collection.

The median Ct values did not differ across the different variants detected (Kruskal-Wallis, *p* = 0.53) or by participant age groups (Kruskal-Wallis, *p* = 0.35; **S2 Fig**). Based on the community surveillance platform, there was no difference in symptom status across the three subvariants (JN.1-like, XBB.2.3-like and XBB.1-like) infected participants (*p* = 0.31). Pairs of first and subsequent episode viruses were sequenced in six of the seven suspect infection cases. Notably, in all cases except one, the lineage in the subsequent virus was found to be the same although there was multiple nucleotide differences detected (**see below**). Samples from both the first and second episodes recovered the KT.1 sequence in three participants, JN.1 in one participant, and JN.1.4.7 in one participant. Only a single participant exhibited a different lineage in the second episode, where the first infection was with JN.1.4.7 and the second with KH.1 (**Table 2**).

### Divergence of the detected XBB.2.3-like and JN.1-lineages

Most (74.1%) of the XBB.2.3-like detections in the 2023/24 KHDSS wave had the GE.1.2 lineage (alias XBB.2.3.10.1.2) backbone, which is characterised by the amino acid changes spike (S): A376S and S:X478T. Three descendants of this lineage were observed: KT.1 (S: K77R), KT.1.2 (ORF1b: N232S) and GE.1.2.2 (S: S408N, ORF1b: A517V, ORF1b: D1899E, ORF7a-ORF8 completed deleted). These were detected in both the community-based and outpatient surveillance platforms. Globally, the GE.1.2 lineage was first detected in the USA (February 4, 2023). Detections in Kenya started in May 2023, mostly in Nairobi (**Fig 2**) ^23^. The first detection of GE.1.2 in Kilifi was in November 2023, although this virus had acquired additional non-synonymous (ORF1a: Q1519H, ORF3a: L52F, ORF3a: W128C and ORF6: D61H) and synonymous mutations (T2506C, T25345C, 26340T; **S3 Fig)**. The Kilifi KH.1 genomes clustered separately from the rest of the available global KH.1 sequences (**S4 Fig**). Within the XBB.2.3-like lineages there was varied within lineage genetic diversity the median pairwise nucleotide differences ranging from 7 to 22 (**S5 Fig**). The KH.1 lineage showed a mean evolutionary rate (substitutions/site/year) of 4.5 x 10^−4^ (95% highest posterior density (HPD)1.3 x 10^−4^ - 8.2 x 10^−4^) while the other XBB.2.3-like lineages was 8.0 x 10^−4^ (95% highest posterior density (HPD) 5.1 x 10^−4^ – 1.0 x 10^−3^).

Most of our JN.1-like detections had the JN.1 lineage (alias BA.2.86.1.1) backbone characterised by amino acid changes S: L455S, ORF1a: R3821K, ORF7b: F19L. Six descendants of this lineage were observed: JN.1.1 (ORF1a: F499L), JN.1.4 (ORF1a: T170I), JN.1.4.7 (ORF3a: G18D), LE.1 (S: R346T), JN.1.10 (S: T95I), and JN.1.16.3 (S: T572I). Within the JN.1.4.7 lineage, we observed clustering per geographic region among the Kenya sequences (**S4 Fig**). Two of the JN.1.4.7 sequences from Kilifi had a ORF1a: T1881I mutation that was lacking in the Nairobi sequences. We observed within lineage genetic diversity among the JN.1-like lineages with a median pairwise nucleotide difference ranging from 3 in JN.1.4.7 to 44 in JN.1.4 **(S5 Fig)**. The estimated evolutionary rate for the JN.1-like variants was 9.8 x 10^−4^ (95% HPD 5.6 x 10^−4^ - 1.3 x 10^−3^) (**S5 Table**). The sequences from the three platforms were interspersed in lineage-specific trees, indicating similar virus sequences identified in the community-based surveillance and among those seeking medical care, whether as outpatients or inpatients.

### Sequence relatedness in suspected reinfection cases

The sequence data from the suspected reinfection cases were compared through pairwise nucleotide comparisons to the earliest sequenced sample from each reinfected individual (**S1 Fig**). Note that, due to the age cut-off for COVID-19 vaccination in Kenya (over 12 years), all suspected reinfection cases listed here were ineligible to receive the COVID-19 vaccine, except for one individual aged between 20 and 29 years. The vaccine information for this individual is missing. As expected, the individual infected by two different Omicron subvariants (JN.1.4.7 then KH.1) exhibited the highest pairwise genetic distance (88 nt). Consistently also, samples collected within a two-week period (<14 days) showed minimal nucleotide differences (<3 nt). In contrast, all except one of the individuals who tested positive again after the two-week window displayed considerable genetic variation in the later samples more consistent with a reinfection (**Table 2**). Notably, in four participants who appeared reinfected by the same Omicron lineage, samples collected after the two-week period exhibited nucleotide differences of 4 nt, 17 nt, 27 nt, and 29 nt, indicating probable reinfections with strains within the same Pango lineage. The one exception was a participant (0.68 years old) who appeared to have been reinfected 25 days later with a genetically identical virus as the first infection **(S1 Fig)**.

### Dynamics of virus introduction and exportation events to and from KHDSS

Using the mugration approach ^24^, we inferred patterns of virus introduction and exportation events into and out of the Kilifi HDSS area. We first reconstructed time-resolved maximum likelihood phylogenies for the XBB.1-like, XBB.2.3-like and JN.1-like lineages based on the 185 newly sequenced Kilifi SARS-CoV-2 genomes and 18,728 “background” genomes from other countries and the Nairobi region in Kenya (**Fig 3**). From these time-resolved phylogenies, we estimated at least 38 SARS-CoV-2 introduction events into Kilifi, 22 for JN.1-like, 14 for XBB.2.3-like, and two for XBB.1-like. The introductions were predominantly from North America (*n* =19), Europe (*n* =18), and Nairobi-Kenya (*n* =1). We also identified 18 exportation events from Kilifi (a) to other regions in Kenya (*n* =4) and (b) to countries in Europe (*n* =7), Asia (*n* =3), North America (*n* =2), Oceania (*n* =1) and Africa (*n* =1). Of these export events, nine involved JN.1-like lineages while six involved for XBB.2.3-like lineages (**Fig 3**).

**Fig 3:**
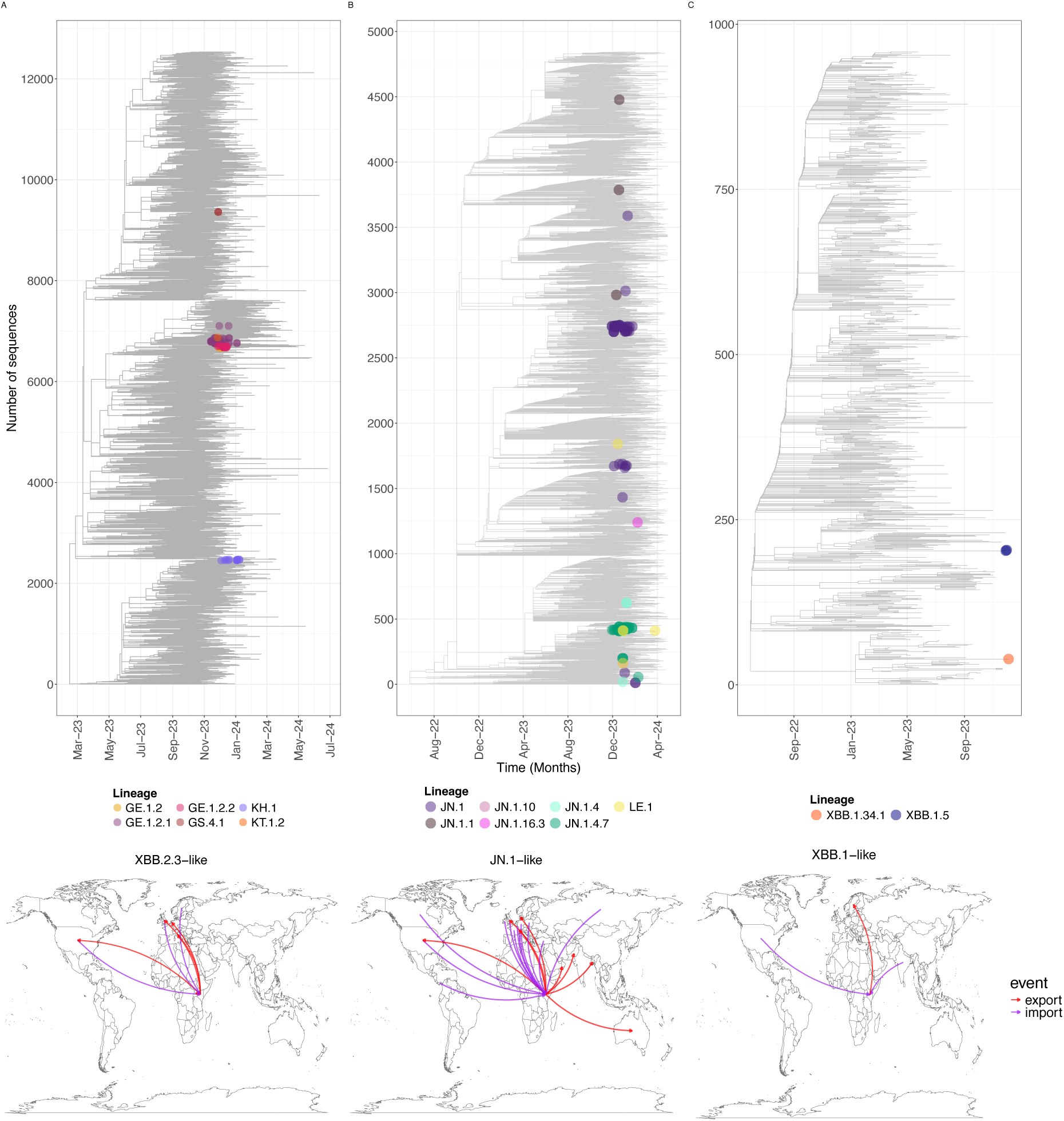
)Top) Time-resolved phylogenetic trees for XBB.2.3-like (**A**), JN.1-like (**B**), and XBB.1-like (**C**) lineages, including sequences sampled on a global basis. The tip points are Kilifi sequences colored by the Pangolin lineages and the non-colored tips are global sequences for context. (Bottom) Global patterns of introduction and exportation events across the observed XBB.2.3-like, JN.1-like and XBB.1-likevariants using time-resolved phylogenies. The arrows move from country of origin to destination are colored by the importation (blue) and exportation events (green) to and from coastal Kenya

## Discussion

In the absence of adequate continued SARS-CoV-2 genomic surveillance in LMICs, we have maintained the genomic surveillance and reported the epidemiology and viral genomic dynamics of SARS-CoV-2 wave between November 2023 and March 2024 in the KHDSS area of Kilifi, Kenya. Genomic analysis of over 180 samples revealed that the wave was dominated by the GE.1.2. * lineage, an XBB.2.3-like sub-variant, and JN.1-like lineages that arrived through direct or indirect multiple virus introductions from outside Kenya. Limited cases of XBB.1.5 and XBB.1.34 were identified but did not appear to spread widely. Our analyses provide the first detailed description of the introduction and further dissemination of JN.1*, XBB.1* and XBB.2.3* sub-variants in Kenya.

The XBB.2.3* strains are a descendent of the recombinant XBB variant ^25^, and XBB evolution resulted in mutations enhancing its immune escape relative to earlier Omicron lineages [8] ^27^. Among these is XBB.2.3* (characterised by S: D253G and S: P521S) that we report here, and which emerged in later in 2022 ^28^. Available data on public sequence databases (e.g., GISAID ^29^) suggests that XBB.2.3* spread worldwide. Its prevalence in the USA in May 2023 was ∼3.2% ^30^. The earliest XBB.2.3* available from Nairobi Kenya was collected in May 2023 but the strain was not detected in Kilifi until in November 2023.

The JN.1* lineages are descendants BA.2.86 Omicron sub-variant (first identified July 2023) ^7^. Approximately 62% of the SARS-CoV-2 sequences deposited on GISAID database during our study period were JN.1-like, indicating this subvariant’s global predominance ^29^. The epidemiological success of JN.1* is hypothesised to be in part due to its additional spike amino acid change (L455S) that supports greater antibody escape, greater ACE-2 binding affinity and increased infectivity ^7,31^. However, unlike most regions globally where JN.1-like lineages were predominant in late 2023/early 2024, XBB.2.3-like lineages were dominant in coastal Kenya, comprising 58.3% (95% CI: 50.9%-65.4%) of our sequenced samples.

During our study period several XBB.1.9 descendants (EG.5, HK.3, and HV.1) were reported globally, albeit at low frequency (1-11%) ^29^. These were not detected in our surveillance. Only a single case of XBB.1.5 (S: G252V and S: F486P) was detected during the Kenyan wave, although this lineage had predominated infections in some global locations including the US ^32^. In early in 2023, the FY.4 lineage dominated in the KHDSS area and is thought to have emerged in-country ^15,33^. Overall, therefore, it appears that the dominant virus lineages in Kilifi during new waves of infection are not always aligned with those that dominate globally. This may reflect stochastic founder events or a unique local SARS-CoV-2 immunological landscape that plays a role in selecting locally predominant variants.

In this study, ∼83% of the infected individuals remained asymptomatic based on our community-based surveillance that tests individuals regardless of symptom status. This aligns with our previous findings in the region: 79% (95% CI: 66%-92%) asymptomatic infection among secondary household infection cases ^34^ and 92.4% among individuals received a test for various reasons early in the pandemic ^35^.The high asymptomatic infection prevalence in many Africa settings has been hypothesized to be potentially due to local climate, human genetics, cross-reacting immunity from endemic infections, and potential underreporting of symptoms ^36^. Other regions of the world have reported lower asymptomatic infection rates e.g., ∼6%-47% in China and USA among SARS-COV-2 cases during the Omicron waves^37,38^.

By October 2023, Coastal Kenya had experienced up to seven waves of SARS-CoV-2 infections ^11–13,39,40^. A year earlier (November 2022), SARS-CoV-2 seroprevalence in the region was estimated at 77.4% ^9^ comparable with other regions globally where seroprevalence was estimated to be 50% - 98% during the same period ^41–45^. This high seroprevalence is hypothesized to primarily result from natural infections, as only about 27% of Kenyan adults had received COVID-19 vaccines ^46^. Most of the infections in the wave we describe here were in children (median age 14 years). Adult infections during this COVID-19 wave were lower likely because most adults have some level of immunity from past exposure or vaccination. Notably, Kenya has not recommended COVID-19 vaccinations for children aged under twelve years. Further investigation into the COVID-19 burden among children and the benefits of vaccinating this frequently immunologically naive group is essential.

Omicron variant infections have been associated with milder symptoms compared to earlier variants ^1,4,5,47^. Data on clinical differences among the emergent Omicron lineages is currently limited. A study from Singapore found no significant difference in hospitalization risk among XBB variants but noted a higher risk of severe outcomes associated with XBB.1.16 ^48^. We found similar clinical characteristics across Omicron sub-variants XBB.2.3-, JN.1-, and XBB.1-like. Notably, individuals attending outpatient clinics commonly presented with upper respiratory symptoms (cough and sore throat) and non-respiratory symptoms (fever and headaches) like what was experienced with earlier Omicron lineages ^49^.

While SARS-CoV-2 reinfections have been widely documented globally, particularly following the emergence of the Omicron variant ^50^ data on these cases in sub-Saharan Africa remain limited ^51^. This study is the first to compare genetic sequences from suspected reinfections in the region, providing crucial genomic evidence of repeat infections, some occurring within remarkably short intervals of less than three weeks, with minimal genetic variation among the viruses. Although uncommon, we show that individuals can experience multiple SARS-CoV-2 episodes within the same wave. One study in Spain found that 42% of 26 patients with COVID-19 episodes separated by 20-45 days had reinfections with different variants^52^. Interestingly, these early reinfections showed no specific clinical patterns, occurring predominantly among unvaccinated individuals and those under 18 years of age ^52^. The mechanisms underlying these short-interval reinfections are not fully understood, and immunological characterization of the reinfected participants in our community-based surveillance cohort is currently ongoing.

This study had several limitations. First, genome sequencing of positive samples failed in 23.2% of the samples, largely due to low viral loads. This may impact the lineage proportions observed if some failures were associated with the genetic characteristics of infecting lineage. Second, the health facility (HF) surveillance platform was interrupted for two weeks (Christmas and New Year’s Eve weeks). This interruption period unfortunately coincided with the peak weeks for the epidemic as inferred from the community surveillance. This resulted in missing data on the lineage distribution for the outpatient surveillance at the epidemic peak and clinical presentation. Third, our surveillance is focused on the KHDSS area (∼900 km^2^) with the community surveillance undertaken in the northern area alone. As much as this might be representative of Kilifi, it may be less so for the rest of Kenya. Inclusion of publicly available sequence data from Nairobi identified seven lineages not detected in Kilifi. There is need for a systematic countrywide surveillance platform.

In conclusion, we provide insights into the molecular epidemiology of a SARS-CoV-2 infection wave that occurred from November 2023 to March 2024 in coastal Kenya, during a post-pandemic wave. This is the first detailed description of JN.1 lineage introduction into Kenya and its transmission and evolution in a sub-Saharan Africa setting ^23,53–56^. We found that this (eighth) wave of SARS-CoV-2 infections in Coastal Kenya was comprised of multiple co-circulating lineages within three sub-variants (JN.1*, XBB.2.3*, and XBB.1*), which were seeded several times into the local population. Unlike JN.1 which was most likely first introduced into Kenya shortly before detection in Kilifi, XBB.2.3* lineages were in circulation elsewhere in Kenya since early 2023 although we nonetheless also observed important genetic differences in the Kilifi strains from those earlier detected elsewhere in the country. As was the case for previous waves, most of the SARS-CoV-2 infections during this new wave (>80%) were asymptomatic, although there were still some instances of severe disease. We recommend further studies to dissect the factors responsible for this. This eight wave of infections in coastal Kenya, provides more evidence that SARS-CoV-2 continues to transmit and evolve in humans causing periodic outbreaks in communities. Sustained genomic surveillance informs us of any changes in the local epidemiology of the virus as well as the origins of new waves, allowing optimisation of available countermeasures.

## Methods

### Study design and population

This study was conducted within the KHDSS, in the Kilifi County of coastal Kenya (**Fig 1**). The study participants were primarily residents of the KHDSS ^19^ which is run by KWTRP. The KHDSS area covers ∼900 km^2^, has a population size ∼300,000 people of diverse ethnic groups with the predominant communities being Mijikenda-speaking people ^19^. The population is characterized by a youthful demographic profile and includes both rural and urban settings.

Respiratory specimens were collected from individuals recruited across three respiratory disease surveillance platforms established by the Pathogen Epidemiology and Omics (PEO) Group of KWTRP. The platforms include:

a. Paediatric inpatient (IP) pneumonia surveillance: since 2002, respiratory samples (nasal washing, nasopharyngeal aspirates, or NP/OP swabs) are collected from children aged <5-year-old admitted to Kilifi County Referral Hospital (KCRH) presenting with symptoms defined as severe or very severe pneumonia according to the WHO definition ^57^. Note that although KCRH serves both residents and non-residents of KHDSS, most of patients seeking care at KCRH are KHDSS residents.
b. Health facility outpatient (OP) surveillance: since December 2020, NP/OP swabs are collected each week from up to 15 persons (±5; any age) presenting with acute respiratory illness (ARI) to each of the five selected outpatient health facilities within the KHDSS. The selected facilities are Pingilikani, Mavueni, Mtondia, Matsangoni and KCRH outpatient department ^20^ (**Fig 1**).
c. Homestead community-based surveillance: this is through the <u>res</u>piratory <u>vi</u>rus <u>re</u>infections (ResViRe) study that started on August 31, 2023, across five administrative locations: Tezo, Roka, Zowerani, Matsangoni, and Ngerenya. Here, participants are visited weekly at home and their respiratory disease status, travel history and NP/OP swabs collected from all recruited homestead members regardless of symptom status. If one or more homestead member(s) is identified positive with SARS-CoV-2, RSV or influenza A/B, sample collection frequency for the homestead is increased to twice a week until all members test negative. By March 31^st^, 2024, 64 homesteads (658 participants) had been enrolled and sampled at least once.

For both IP and OP surveillances, samples and relevant participant demographic details (e.g., sex, date of birth, COVID-19 vaccination status, and location/facility) and clinical data were collected once during participant contact with the hospital or health facility where they were recruited ^20^. Clinical illness signs and symptoms were recorded by the attending clinician based on a standard questionnaire. The following symptoms were recorded following interview with participant or their caregiver: cough, headache, fever (above 37.8°C), sore throat, vomiting, diarrhoea, difficulty breathing, wheezing, nausea, joint pains and chest pains.

For the ResViRe study, participant baseline information was captured at recruitment. This included age, sex, vaccinations received, co-morbidities, social economic status, illnesses in the last week among others. During weekly home visits, a trained fieldworker administered short questionnaire to collect information about participant current respiratory wellness status, any illness that may have occurred since the last visit, recent travel and any contact with the healthcare system.

### Laboratory methods

#### Nucleic acid extraction and SARS-CoV-2 screening

Ribonucleic acid (RNA) was extracted from 140μl NP/OP samples using the automated RNeasy Mini Kit (QIAGEN, UK) following the manufacturer’s protocol. SARS-CoV-2 was screened using an in-house real-time RT-PCR with primers/probe targeting the envelope (E) gene (forward: 5’-ACA GGT ACG TTA ATA GTT AAT AGC GT -3’, reverse: 5’-ATA TTG CAG CAG TAC GCA CAC A -3’ and probe 5’-ACA CTA GCC ATC CTT ACT GCG CTT CG-3’) and QIAGEN Multiplex RT-PCR + R Kit (QIAGEN, UK). Negative and positive controls starting from both the extraction and RT-PCR stages were included in PCR plate. A sample was considered positive if it had a cycle threshold (Ct) of ≤35.0.

#### SARS-CoV-2 whole genome sequencing (WGS)

Positive SARS-CoV-2 samples were subjected for WGS. RNA was re-extracted from 140µl of sample using the QIAamp Viral RNA Mini Kit (QIAGEN, UK), reverse-transcribed using LunaScript RT SuperMix Kit (New England Biolabs, UK), and the cDNA PCR amplified using Q5 Hot Start High-Fidelity 2x Mastermix (New England Biolabs, UK) with the ARTIC nCoV-2019 version 5.3.2 primers ^58^. Briefly, 8ul of RNA and 2μl of LunaScript RT Mix (NEB, E3010, MA, USA) was incubated at 25◦C for 2 min, 55 °C for 10 min, 95 °C for 10 min then held at 4 °C. The cDNA was amplified in 2 reaction mixes per sample with the ARTIC nCoV-2019 version 5.3.2 primer pools A and B. The amplification reactions were set up by combining 6.3 μl of Q5 Hot Start High-Fidelity 2X Master Mix (NEB M0494, MA, USA), 1.9 μl of nuclease-free water, 2 μl of primer pool and 1.3 μl of cDNA. The thermocycling conditions were: 1 cycle of 98°C for 30 s, followed by 25 cycles of 98 °C for 30 s and 65 °C for 5 min, 15 cycles of 62.5 °C for 5 min and 98 °C for 15 s, 1 cycle of 62.5 °C for 5 min and held at 4 °C indefinitely. The two reactions were pooled and cleaned using 1X AMPure XP beads (Beckman Coulter, A63881, Indianapolis, USA) as per the manufacturer’s instructions. The cleaned amplicons were eluted in 20ul and quantified using a QUBIT fluorometer. Samples with a concentration of > 18 ng/μl were taken forward for library preparation. Sequencing library preparation was undertaken using the COVIDSeq assay (Illumina, US) and sequenced on the Illumina Miseq platform to generate 100bp paired end reads while multiplexing 80 to 100 samples per run.

### Bioinformatic data analysis

#### Genome assemblies and consensus generation

Consensus genomes/sequences were generated by mapping the quality filtered (using fastp v.0.23.4; q≥30 and adapters removed) fastq reads to the Wuhan reference (MN908947.3) using bwa v0.7.17, sorting the mapped reads using samtools v1.10, removing primer sequences and calling the consensus with a minimum depth of 10 reads using ivar v1.4.2 as previously described ^39^.

#### Quality control, lineage and clade assignment

The quality of the newly assembled sequences was assessed using web Nextclade program version 2.14.1 ^59^. Key parameters evaluated included no sequence recovered from NC, expected sequence for the PC. Samples yielding complete or near complete genomes sequences (>70% coverage) were classified into SARS-CoV-2 lineages using the Phylogenetic Assignment of Named Global outbreak Lineages (Pangolin software v1.28) and Nextclade v3.7.1 tools ^59^. For the serial samples collected from same individual in the ResViRe study, only one genome was selected per infection episode (see definition below) and included into downstream analysis based on genome completeness.

#### Global and other Kenya comparison data (“background data”)

This was done per major lineage detected (i.e., XBB.2.3*, JN.1*, XBB.1*). The spatial and temporal distribution of the global comparison data is summarised in **S6 Fig**. For XBB.2.3-like lineages, all genomic data (*n* = 12,576) on GISAID ^29^ from 1^st^ October 1, 2023, to June 30, 2024, were downloaded and used for the subsequent phylogenetic analyses. For the JN.1-like lineages, over 500,000 sequences collected between October 1st, 2023, to April 30^th^, 2024, were available on GISAID. Metadata for all the sequences was downloaded and subsampled in R to include five near-complete sequences per country, month of collection and lineages identified. This resulted in a total of 5,230 sequences used for downstream phylogenetic analysis. For XBB.1-like lineages, 103 XBB.1.34.1 sequences and 187,083 XBB.1.5 sequences were available on GISAID by October 1, 2024. The metadata was downloaded and the XBB.1.5 sequences downsampled to include 10 sequences per country and month of collections to give 920 sequences from August 1, 2022, to November 30, 2023. All the 103 XBB.1.34.1 sequences were included.

All Kenyan SARS-CoV-2 genomic data and metadata from Jan 1, 2023, to October 1, 2024, was downloaded from GISAID and included in the analysis to describe the genomic epidemiology of the virus within the country. The genomic data included 775 sequences collected from 19 counties in Kenya and was classified into 69 pango lineages. The spatial and temporal distribution of the data is shown in **Fig 2**.

All genome sequences and associated metadata in the global dataset are published in GISAID’s EpiCoV database and can be accessed at https://doi.org/10.55876/gis8.241022hw. The EPI_SET_241022hw contains data from 111 countries and territories

#### Pairwise nucleotide differences

Pairwise nucleotide differences from sequence alignments for each lineage were calculated by iterating through all possible sequence pairs using a custom Python script that utilizes Biopython (available on https://github.com/arnoldlambisia/Epiinsights_wave8_Kilifi.git). Only valid sites i.e., A, C, T and G were considered.

#### Phylogenetic analyses

Multiple sequence alignments (MSA) were generated using Nextalign v1.10.1 (https://github.com/neherlab/nextalign) with the Wuhan-Hu-1 SARS-CoV-2 genome (Accession number: NC_045512.2) used as the reference. The alignments were manually checked using AliView v1.27 to determine if any sequences were duplicates or indicative of sequencing/alignment errors and used as input for a maximum likelihood phylogenetic analysis using IQTREE v2.1.3 (http://www.iqtree.org/) assuming the general time reversible (GTR) model of nucleotide substitution. Branch support was evaluated with 1,000 bootstrap replications. Pairwise nucleotide differences were calculated using *pairsnp* programme (https://github.com/gtonkinhill/pairsnp).

### Estimation of molecular clock signal and evolutionary rate

The ML trees were checked for a temporal signal in genetic divergence over time using Tempest v1.5.3 (http://tree.bio.ed.ac.uk/software/tempest/) and outlier sequences dropped. The ML trees were subsequently used to generate time-resolved trees using TreeTime v0.11.1 ^24^.

Estimation of the most recent common ancestor (TMRCA) and evolutionary rate of the identified variants in Kilifi was performed using the Bayesian software BEAST v1.10.4 ^60^. The Kenyan sequences were grouped; JN.1-like (n=87), XBB.2.3-like (n=102, KH.1 removed) and KH.1 (n=35). XBB.1-like were not included due to small numbers. ML trees were generated using IQTREE v2.1.3 (http://www.iqtree.org/) and used to inspect for a temporal clock signal using Tempest v1.5.3 (http://tree.bio.ed.ac.uk/software/tempest/). Time resolved phylogenies were estimated using the uncorrelated relaxed clock, HKY substitution model and the Bayesian skyline model. Analysis was run using 100 million Markov chain Monte Carlo (MCMC) states sampling every 50,000 steps. Tracer v1.71 ^61^ was used to check for convergence (effective sample size > 200).

### Phylogeographic analysis

Virus introductions events into Kilifi for each sub-variant were inferred from a phylogeographic reconstruction based on the Kenyan genomic sequences data and subsampled background global genomic sequences retrieved from GISAID ^29^. These background genomic sequences were selected as described above. Specifically, we conducted a mugration analysis as implemented in the TreeTime program, using the country of origin for the global data and Kilifi and non-Kilifi data for Kenyan data as discrete traits to reconstruct geographic locations at all internal nodes. A custom Python script (available on https://github.com/arnoldlambisia/Epiinsights_wave8_Kilifi.git) was then used to infer the number of transition events using the annotated tree from the mugration analysis by traversing the tree from root to the most external tip and counting every base change and record the timing of that change ^62^.

#### Mutational spectrum

To identify evidence of local evolution, nucleotide and amino acid mutations across the sequences were visualized using Snipit v1.4 (https://github.com/aineniamh/snipit) against a local XBB.2.3-like, and JN.1-like reference. The potential fitness effects of the lineage defining mutations were checked using the https://jbloomlab.github.io/SARS2-mut-fitness/tool (**S3 table**) ^63^.

### Epidemiological data analyses

#### Definitions

a. Symptomatic infection was defined as occurrence of one or more respiratory or systemic infection symptoms (listed in the next section) at sample collection or during the preceding or subsequent week after sample collection for the community cases. The following symptoms were recorded following interview with participant or their caregiver: cough, headache, fever (above 37.8°C), sore throat, vomiting, diarrhoea, difficulty breathing, wheezing, nausea, joint pains and chest pains.
b. Symptomatic fraction was calculated as the proportion of infected individuals who developed/reported having symptoms among the community-study (ResViRe) participants.
c. Infection episode was defined as a period in which a study participant provided an NP/OP sample that were positive for SARS-CoV-2 by qRT-PCR, this sandwiched by negative RT-PCR test in both preceding and subsequent two weeks.
d. An introduction event to the Kilifi, Kenya was defined as a lineage transition event inferred by the mugration analysis from any other country or Kenya region outside Kilifi to Kilifi, Kenya.
e. A lineage was defined as a SARS-CoV-2 strain designation as described in ^64^ based on a standard questionnaire.

### Statistical analyses

Temporal trends of SARS-CoV-2 positive cases and lineages were summarized either weekly or monthly. Prevalence was calculated as the number of samples positive divided by the total number of samples collected/tested per platform and a 95% confidence interval (CI) given by a Wilson Score method. Kruskal Wallis and Wilcoxon tests were used to compare median viral load across different variants, age groups, platforms and symptom status. The distribution of demographic characteristics, variant/lineages proportions and clinical characteristics were summarized and compared across all the different platforms using a chi-square statistic. Univariate and multivariable logistic regression models were used to examine the association between symptom status (symptomatic vs asymptomatic) and various predictor variables including sex, age (0-4, 5-9, 10-19, 20-39, 40-64 and ≥65 years), vaccination status (unvaccinated or vaccinated) and infecting variant (JN.1-like, XBB.2.3-like and XBB.1-like). Results were reported as unadjusted and adjusted odds ratios (ORs) with 95% CIs. p values less than 0·05 were considered statistically significant. The data was analyzed using R version 4.1.1 and STATA version 15.0 ^65^.

### Ethics statement

Each surveillance platform (community, outpatient, and inpatient) that provided samples analysed here had a dedicated research protocol. The protocols’ consenting and sample collection process were reviewed and approved by KEMRI Scientific Ethics and Research Unit (SERU), Nairobi Kenya (protocol numbers #3178, #3103 and 4724). Samples were collected following consent from a parent or guardian for participants aged <18-year-olds (with assent for children aged between 13–18-year-old). Individual written informed consent was sought for participants aged >18 years.

## Data and code availability

The final consensus genomes from the SARS-CoV-2 samples sequenced in this study have been deposited in the Global Initiative on Sharing all Influenza Data (GISAID) database and can be accessed at https://doi.org/10.55876/gis8.250116pz.

Epidemiological data and scripts for data analysis are available on the Harvard dataverse https://doi.org/10.7910/DVN/BMCJTI.

## Acknowledgements

We thank study participants, parents and guardians who consented participation in the studies, members of the Pathogen Epidemiology and Omics (PEO) Group particularly the field team who collected the clinical samples and metadata and laboratory diagnostics team who processed the samples analysed and presented in this report. We thank Prof. D. James Nokes (retired) for his critical role in the design of the parent studies that provided the analysed samples. We thank Mr. Christopher Nyundo for his assistance in generating the study area maps. We thank all genomic data contributors including authors and their originating labs of the sequence data in GISAID that we included in this research. This working is published with permission from director KEMRI.

