## supplementary tables and figures for "Genomic and clinical epidemiology of SARS-CoV-2 in coastal Kenya: Insights into variant circulation, reinfection, and multiple lineage importations during a post-pandemic wave"

### Supporting information

#### Epidemiological insights from the genomic analyses of the SARS-CoV-2 infection wave in coastal Kenya, 2023-24

### Supplementary tables

**S1 Table:** Determinants of SARS-CoV-2 symptom status among community cases in coastal Kenya

| | Unadjusted OR | P value | Adjusted OR | P value |
|---|---|---|---|---|
| **Sex[1]** | | | | |
| **Male** | 0.45 (0.15 - 1.21) | 0.01 | 0.38 (0.11 - 1.17) | 0.11 |
| **Age group[2]** | | | | |
| 5-9 | 0.45 (0.06 - 2.02) | 0.3 | 0.26 (0.01 - 2.02) | 0.3 |
| 10-19 | 0.35 (0.05 - 1.53) | 0.2 | 0.22 (0.01 - 1.39) | 0.2 |
| 20-39 | 1.16 (0.34 -3.72) | 0.7 | 1.37 (0.35 -5.05) | 0.6 |
| 40-64 | 0.97 (0.26 - 3.22) | 0.9 | 1.50 (0.37 - 5.65) | 0.5 |
| 65+ | 7.57e-08 (NA - 1.54e+72) | 0.9 | 4.07e-08 (NA - 3.63e+123) | 0.9 |
| **Vaccination Status[3]** | | | | |
| Vaccinated | 1.06 e-07 (NA - 5.81e+34) | 1.90E-11 | 2.17e-08 (NA - 2.91e+102) | 0.9 |
| **Variant[4]** | | | | |
| XBB.2.3-like | 0.68 (0.26 - 1.78) | 0.4 | 0.61 (0.21 - 1.72) | 0.35 |
| XBB.1-like | 2.0 e+7 (3.5e-122 - NA) | 0.9 | 4.01e+8 (0.00 - NA) | 0.9 |

[1]Reference group is female
[2]Reference group is 0-4
[3]Reference group is unvaccinated
[4]Reference group is JN.1-like

**S2 Table:** Comparison of demographic and clinical characteristics between sequenced and not sequenced samples.

| | Sequenced (*n* = 185) | Not sequenced (*n* = 56) | Total (*n* = 241) | p-value |
|---|---|---|---|---|
| **Platform** | | | | 0.177 |
| Community | 115 (62.2) | 29 (51.8) | 144 (59.8) | |
| Outpatient | 65 (35.1) | 23 (41.1) | 88 (36.5) | |
| Inpatient | 5 (2.7) | 4 (7.1) | 9 (3.7) | |
| **Sex** | | | | 0.424 |
| Female | 107 (57.8) | 29 (51.8) | 136 (56.4) | |
| **Age (Years)** | | | | 0.216 |
| Median (range) | 13 (0.0 - 104.0) | 14 (0.0 - 65.0) | 14 (0.0 - 104.0) | |
| **Age group (Years)** | | | | 0.653 |
| 0-4 | 58 (31.4) | 17 (30.4) | 75 (31.1) | |
| 5-9 | 18 (9.7) | 6 (10.7) | 24 (10.0) | |
| 10-19 | 32 (17.3) | 12 (21.4) | 44 (18.3) | |
| 20-39 | 42 (22.7) | 15 (26.8) | 57 (23.7) | |
| 40-64 | 21 (11.4) | 5 (8.9) | 26 (10.8) | |
| 65+ | 14 (7.6) | 1 (1.8) | 15 (6.2) | |
| **Symptom status** | | | | 0.787 |
| Asymptomatic | 93 (50.3) | 27 (48.2) | 120 (49.8) | |
| Symptomatic | 92 (49.7) | 29 (51.8) | 121 (50.2) | |
| **Clinical characteristics** | | | | 0.932 |
| Coughing | 88 (47.6) | 27 (48.2) | 115 (47.7) | |
| Diarrhea | 3 (1.6) | 0 (0.0) | 3 (1.2) | 0.338 |
| Headache | 15 (8.3) | 1 (1.9) | 16 (6.9) | 0.108 |
| Fever | 44 (23.8) | 15 (26.8) | 59 (24.5) | 0.647 |
| Vomiting | 4 (2.2) | 1 (1.8) | 5 (2.1) | 0.863 |
| Sore throat | 11 (6.1) | 3 (5.8) | 14 (6.0) | 0.927 |
| Chest pains | 5 (2.8) | 1 (1.9) | 6 (2.6) | 0.732 |
| Difficulty breathing | 7 (3.8) | 7 (12.5) | 14 (5.8) | 0.015 |

| | | | | |
|---|---|---|---|---|
| Joint pains | 4 (2.2) | 0 (0.0) | 4 (1.7) | 0.278 |
| Wheezing | 2 (1.1) | 1 (1.8) | 3 (1.2) | 0.677 |

**S3 Table:** Description of PANGO lineages identified in Kilifi, 2023-24.

| Pango lineage | Alias | Frequency (%; *n* = 185) | Lineage defining mutations | Comment (known Impact of changes) |
|---|---|---|---|---|
| **JN.1-like** | | **75 (31.1%)** | **S: L455S, ORF1a: R3821K, ORF7b: F19L** | Increased neutralization resistance [1,2] |
| JN.1 | BA.2.86.1.1 | 33 (13.7) | S: L455S, ORF1a: R3821K, ORF7b: F19 | |
| JN.1.4.7 | BA.2.86.1.1.4.7 | 30 (12.4) | ORF3a: G18D | |
| LE.1 | BA.2.86.1.1.4.7.1 | 5 (2.1) | S: R346T | |
| JN.1.4 | BA.2.86.1.1.4 | 3 (1.2) | ORF1a: T170I | |
| JN.1.1 | BA.2.86.1.1.1 | 2 (0.8) | ORF1a: F499L, C11747T | |
| JN.1.10 | BA.2.86.1.1.10 | 1 (0.4) | S: T95I | |
| JN.1.16.3 | BA.2.86.1.1.16.3 | 1 (0.4) | S: T572I | |
| **XBB.2.3-like** | | **108 (44.8)** | **Defined by S: P521S and S: S486P** | |
| KT.1 | XBB.2.3.10.1.2.1.1 | 56 (23.2) | S: K77R | |
| KH.1 | XBB.2.3.3.1.2.1.1.1.1.1 | 25 (10.4) | S: E554K, on N: L13F, ORF1a: R135S | |
| GE.1.2.2 | XBB.2.3.10.1.2.2 | 16 (6.6) | S: S408N, ORF1b: A517V and ORF1b: D1899E | ORFa-ORF8 possibly completely deleted |
| GE.1.2 | XBB.2.3.10.1.2 | 7 (2.9) | S: N148T | Kenya lineage |
| GS.4.1 | XBB.2.3.11.4.1 | 2 (0.8) | S: N185D and ORF3a:G172C | |
| JE.1.1.1 | XBB.2.3.3.1.2.1.1.1.1 | 1 (0.4) | S: Q52R | |
| KT.1.2 | XBB.2.3.10.1.2.1.1.2 | 1 (0.4) | ORF1b: N2328S, C583T | |
| **XBB.1-like** | | **2 (0.8)** | **Defined by S: G252V** | Low fitness change [2] |
| XBB.1.34.1 | | 1 (0.4) | S: E554K, ORF1b: P970 | |
| XBB.1.5 | | 1 (0.4) | S: F486P | |

Description based on information provided in https://github.com/cov-lineages/pango-designation/blob/master/lineage_notes.txt
Note that two lineages are thought to have arisen in Kenya

**S4 Table:** Comparison of demographic characteristics and clinical presentation across SARS-CoV-2 variants among cases in Kilifi.

| | JN.1-like (*n* = 75) | XBB.1-like (*n* = 2) | XBB.2.3-like (*n* = 108) | Total (*n* = 185) | p value |
|---|---|---|---|---|---|
| **Sex** | | | | | 0.171 |
| Female | 41 (54.7) | 0 (0.0) | 66 (61.1) | 107 (57.8) | |
| **Age group in years** | | | | | 0.973 |
| 0-4 | 23 (31.1) | 1 (50.0) | 34 (31.5) | 58 (31.5) | |
| 5-9 | 8 (10.8) | 0 (0.0) | 10 (9.3) | 18 (9.8) | |
| 10-19 | 11 (14.9) | 0 (0.0) | 21 (19.4) | 32 (17.4) | |
| 20-39 | 17 (23.0) | 1 (50.0) | 24 (22.2) | 42 (22.8) | |
| 40-64 | 10 (13.5) | 0 (0.0) | 10 (9.3) | 20 (10.9) | |
| 65+ | 5 (6.8) | 0 (0.0) | 9 (8.3) | 14 (7.6) | |
| Missing data | 1 | 0 | 0 | 1 | |
| **Symptom status** | | | | | 0.307 |
| Asymptomatic | 40 (53.3) | 0 (0.0) | 53 (49.1) | 93 (50.3) | |
| Symptomatic | 35 (46.7) | 2 (100.0) | 55 (50.9) | 92 (49.7) | |
| **Clinical presentation** | | | | | |
| Coughing | 35 (46.7) | 2 (100.0) | 51 (47.2) | 88 (47.6) | 0.327 |
| Fever | 16 (21.3) | 1 (50.0) | 27 (25.0) | 44 (23.8) | 0.578 |
| Headache | 6 (8.2) | 0 (0.0) | 9 (8.6) | 15 (8.3) | 0.909 |
| Sore throat | 5 (6.8) | 0 (0.0) | 6 (5.7) | 11 (6.1) | 0.892 |
| Difficulty breathing | 3 (4.0) | 0 (0.0) | 4 (3.7) | 7 (3.8) | 0.956 |
| Chest pains | 3 (4.1) | 0 (0.0) | 2 (1.9) | 5 (2.8) | 0.659 |
| Vomiting | 3 (4.0) | 0 (0.0) | 1 (0.9) | 4 (2.2) | 0.364 |
| Joint pains | 2 (2.7) | 0 (0.0) | 2 (1.9) | 4 (2.2) | 0.912 |
| Diarrhoea | 2 (2.7) | 0 (0.0) | 1 (0.9) | 3 (1.6) | 0.646 |
| Wheezing | 1 (1.3) | 0 (0.0) | 1 (0.9) | 2 (1.1) | 0.956 |

### Supplementary figures

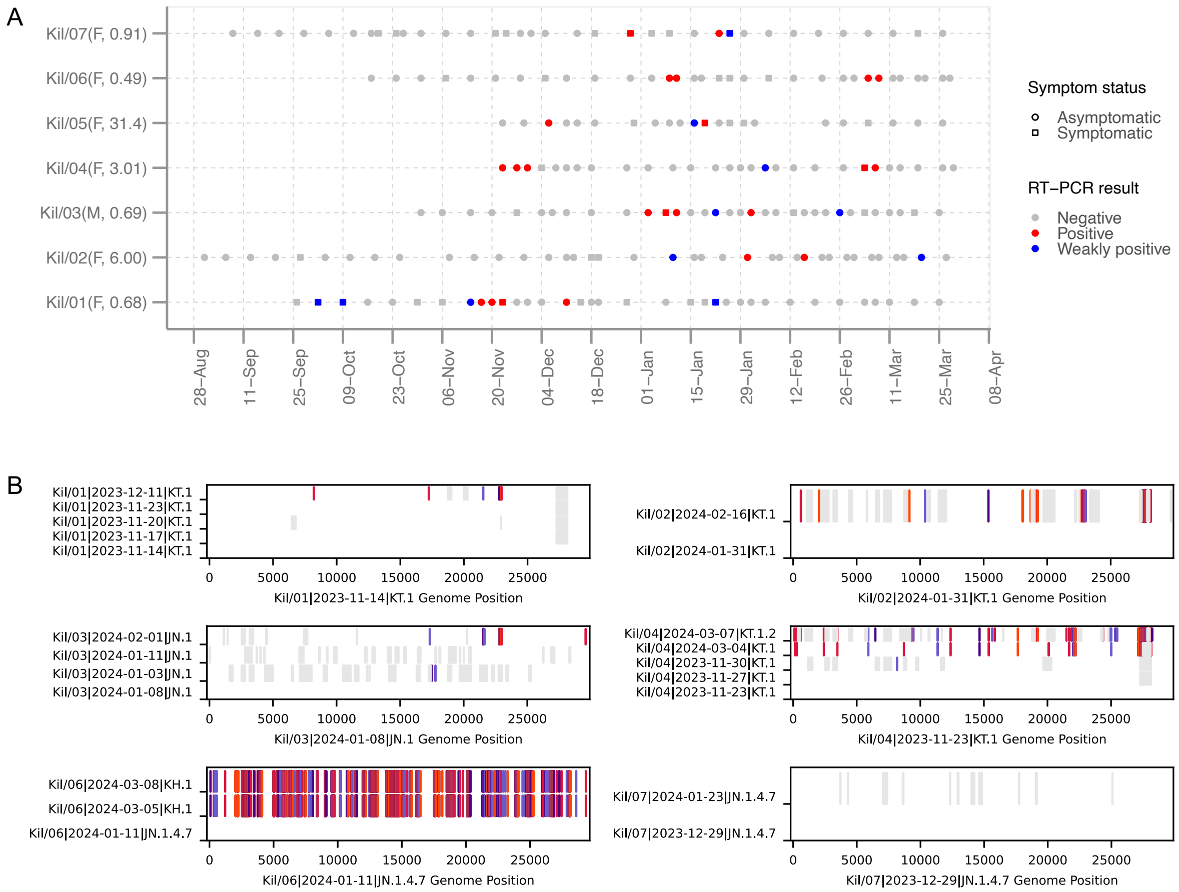

#### S1 Fig: Suspected reinfections in the community-based surveillance and genomic findings.

**(A)** NP/OP temporal sampling patterns in the seven participants with suspected reinfection events The-axis labels show participant number and in brackets their sex (M for male and F for female) followed by their age in years**. (B)** Each panel is a sequence alignment for positive samples from six of the seven suspected reinfection cases. The genomes are compared against the earliest positive sample from the individual (bottom line of each graph) except in Kil/03 where the second sequence was used due to many gaps observed in the initial sample. Colored bars indicate single nucleotide changes from the initial patient sequence with the color coding ...

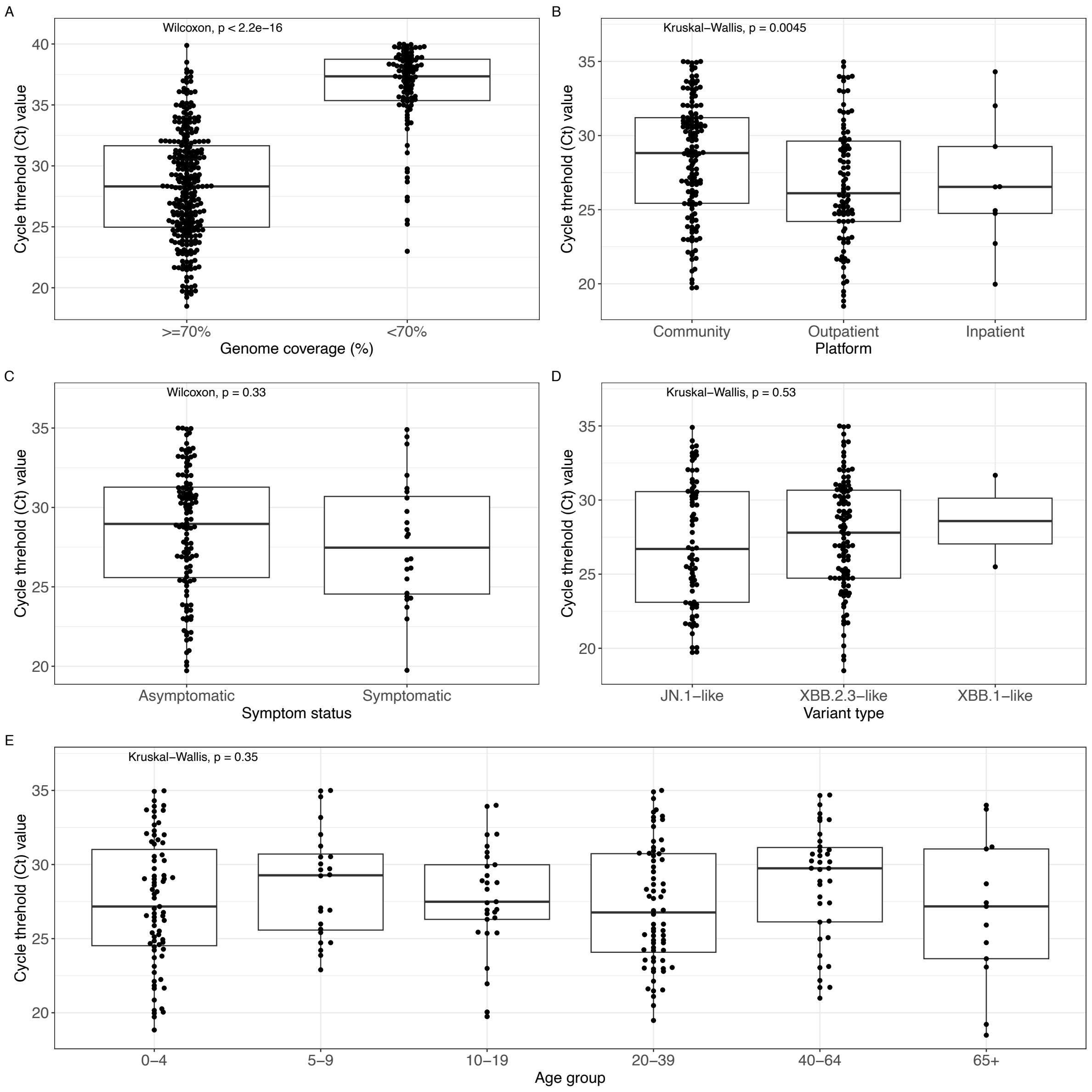

**S2 Fig: Patterns of SARS-CoV-2 cycle threshold (indicative and inverse to virus load quantities).** Comparison across different genome completeness (**A**) surveillance platforms (**B**), clinical presentation (**C**; only for community surveillance), sub-variants (**D**) and age groups (**E**) in samples collected between November 2023 and March 2024 from coastal Kenya.

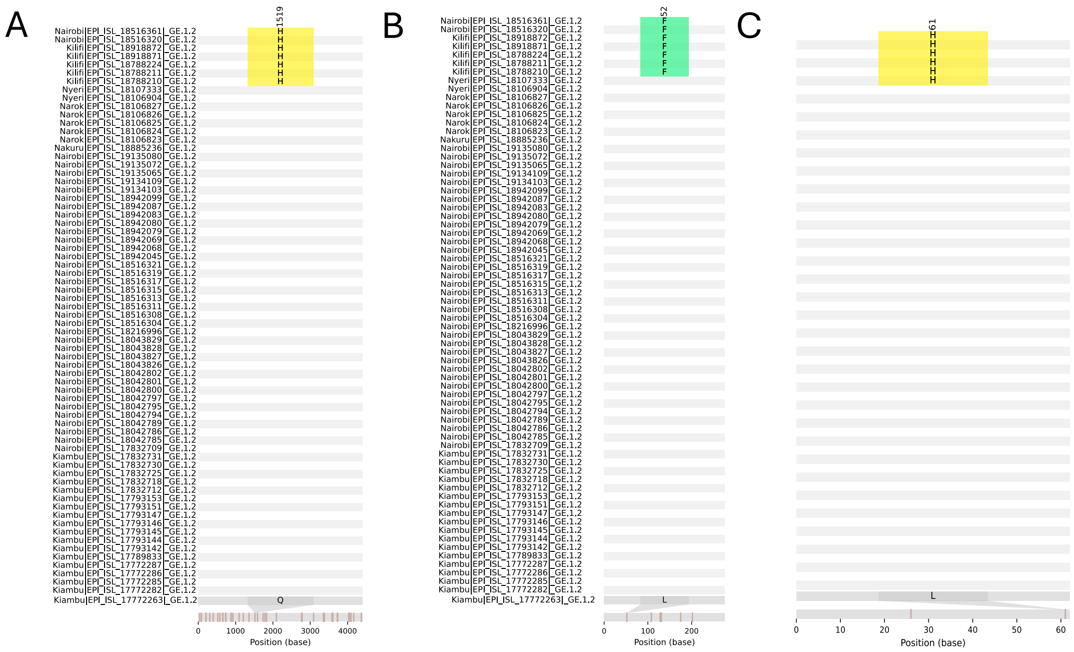

S3 Fig: Amino acid differences in the ORF1a, ORF3a and ORF6 among lineage GE.1.2 sequences (n = 62) collected in multiple locations across Kenya. The changes highlighted in a rectangular box are mutations that were seen mainly in Kilifi sequences.

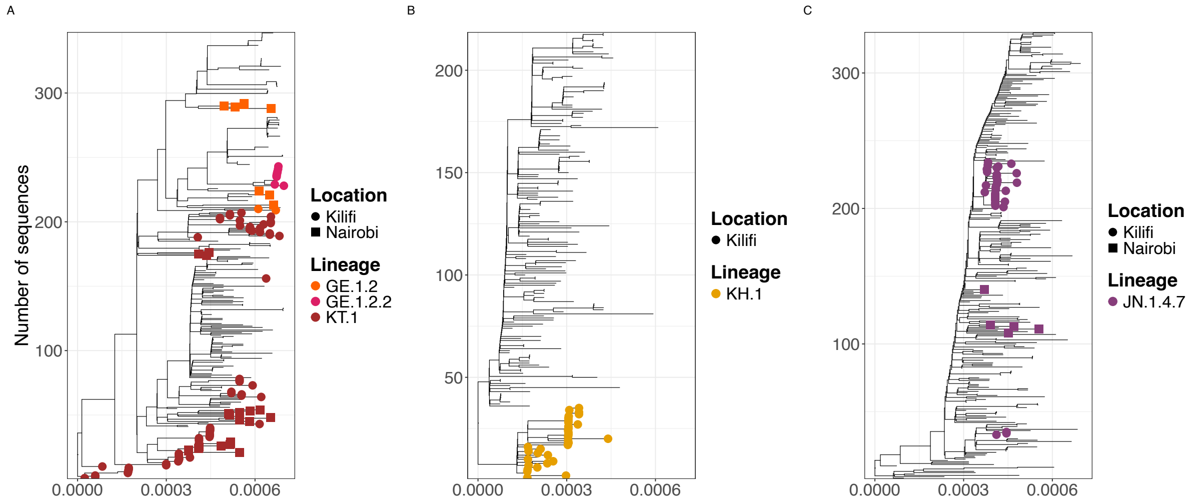

S4 Fig: Maximum likelihood trees for five lineages commonly detected lineages. The phylogenies show the evolutionary clustering of Kenyan and global sequences of the lineages collected between October 2023 and June 2024. Global data for contextual purposes are shown as non-colored tips and Kenyan sequences are colored The tips are shaped by location in Kenya either Nairobi or Kilifi.

.

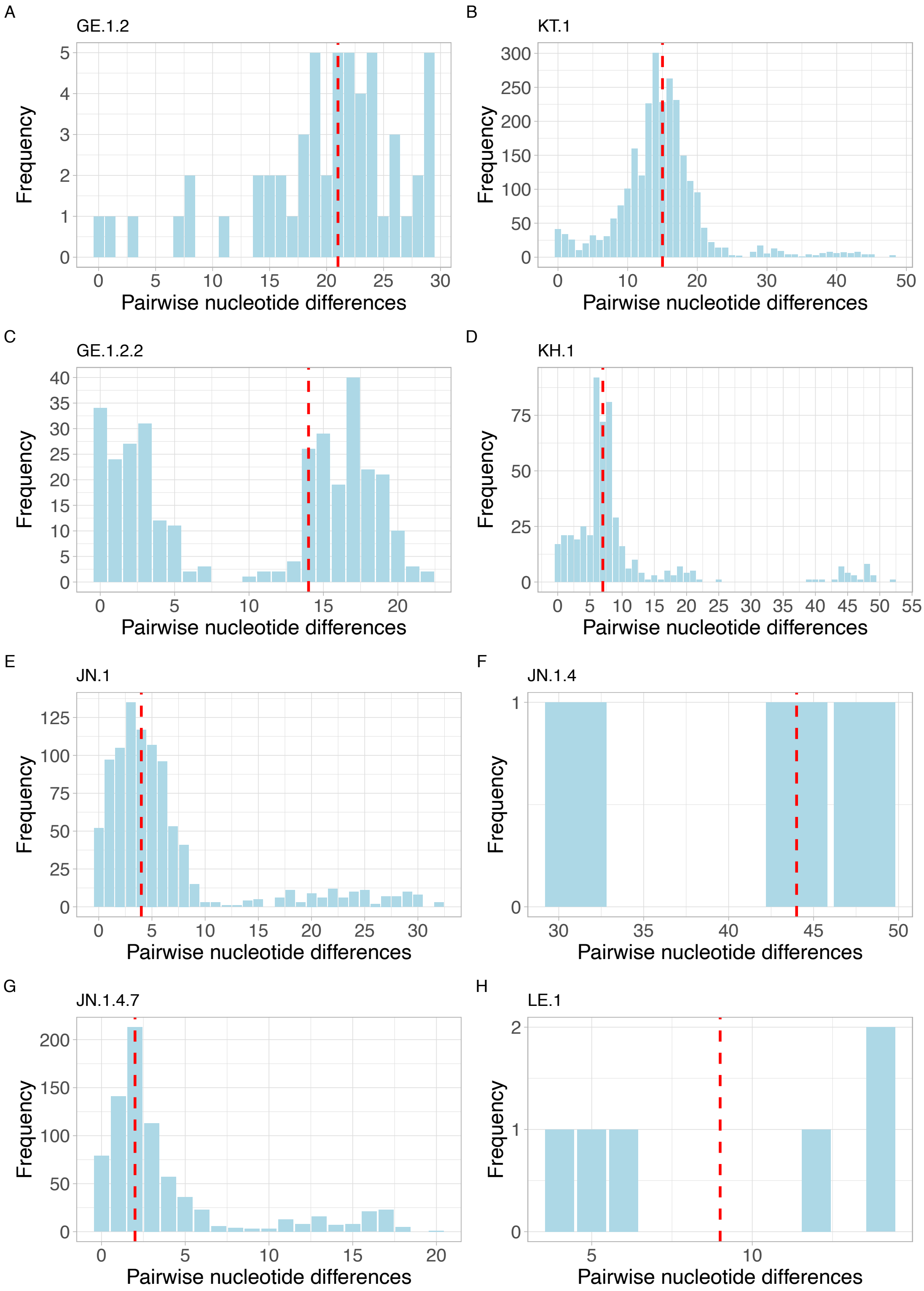

**S5 Fig: Genetic diversity within identified lineages in Kenya between October 2023 and April 2024.** The bar plots show pairwise nucleotide differences between all possible sequence pairs within a lineage. The red dotted line shows the median pairwise nucleotide difference within the lineage.

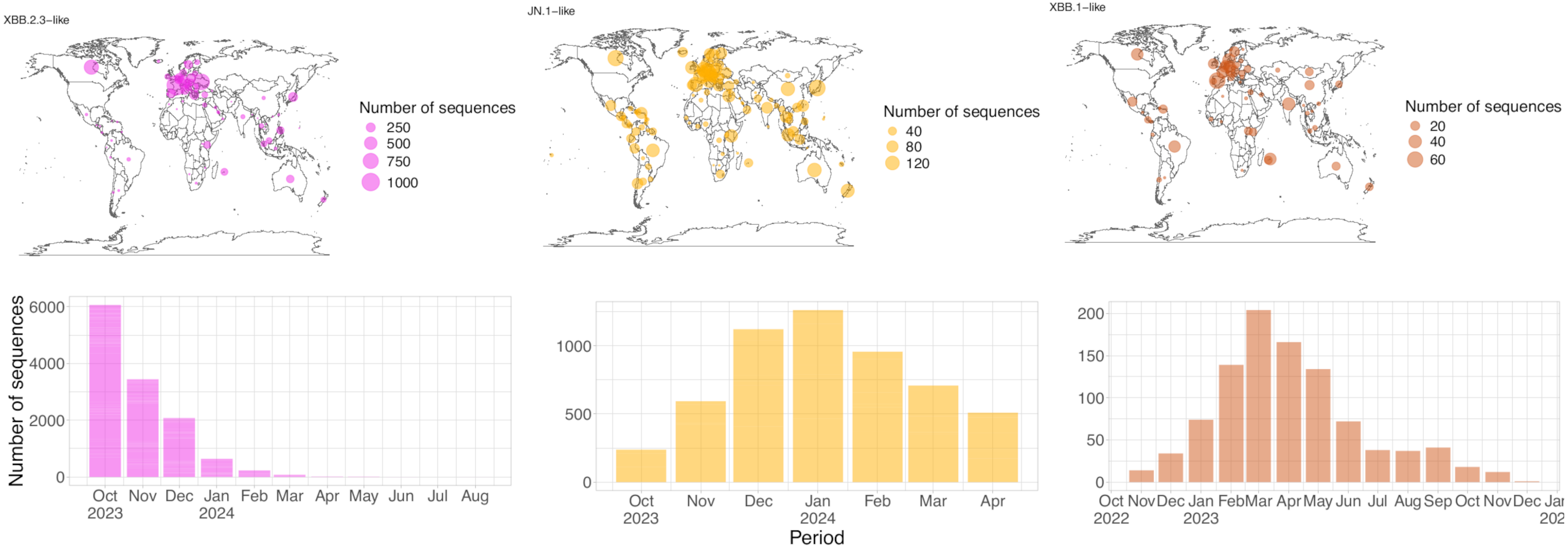

**S6 Fig: Spatial and temporal distribution of sampled SARS-CoV-2 sequence from GISAID for XBB.2.3-like, JN.1-like and XBB.1-like variants.** **(Upper panels)** World maps showing the origin of subsampled sequences with the size of the circles corresponding to the number of sequences for XBB.2.3-like, JN.1-like and XBB.1-like. (**Lower panels**) Temporal distribution (monthly) of the subsampled sequences for XBB.2.3-like, JN.1-like and XBB.1-like subvariants.
